# Machine-Assisted Topic Analysis of Large-Scale Health Experience Data: Identifying Sociodemographic Differences and Evaluating Bias in Large Language Models

**DOI:** 10.64898/2026.05.20.26353755

**Authors:** Paulina Bondaronek, Emma Ward, Emma Beecham, Eric Zhang, Yuqing Huang, Julia Ive, Felix Naughton, Honghan Wu, Cecilia Vindrola-Padros

## Abstract

**Introduction:** Large-scale free-text data with socio-demographic information can capture nuanced accounts of lived experience that are difficult to detect in structured measures. However, manual qualitative analysis is difficult to scale, while automated approaches may obscure subgroup variation or introduce bias. This is especially relevant for large language models (LLMs), whose use in qualitative health research is increasing despite limited evaluation in socio-demographically stratified analysis.

**Objectives:** This study examined how socio-demographic differences in health and wellbeing experiences were manifested in a large-scale free-text dataset, and evaluated how different AI-assisted analytic approaches identified these differences. Specifically, it aimed to: (1) identify socio-demographic differences using Machine-Assisted Topic Analysis (MATA); (2) compare MATA outputs with topic modelling combined with LLM-based topic interpretation; and (3) examine potential bias in LLM-based analysis.

**Methods:** We analysed 2,177 valid free-text responses from the UK COVID-19 Well-being Tracker, a longitudinal survey of adults recruited during the pandemic. Responses described factors influencing health behaviours, mood, and wellbeing over time. Data were preprocessed and stratified by gender, socioeconomic status (SES), age. MATA combined topic modelling, using Latent Dirichlet Allocation, with human-led qualitative interpretation of topic keywords and representative responses. The same topic model outputs were then interpreted using an LLM for comparison. Potential LLM bias was assessed using a demographic label-swap crossover design, with bias evaluated through Jaccard lexical similarity, VADER sentiment, and NRC emotion analysis. Grounded Review and Assessment of Computational Evidence (GRACE) was used to evaluate the AI outputs.

**Results:** MATA identified meaningful socio-demographic thematic differences in pandemic-related mood and wellbeing across gender, SES, and age. Common themes included disruption, adaptation, uncertainty, routine, and the influence of work, relationships, and health on wellbeing. Male-stratified topics emphasised routines, habits, and coping with external pressures, whereas female-stratified topics were more relational and reflective, focusing on connection, isolation, family wellbeing, and anxiety. Lower SES narratives included practical strain, financial pressure, and loss of control, while higher SES narratives more often reflected adjustment, autonomy, and meaning-making. Older adults described health, gratitude, and family connection, whereas younger adults emphasised work-related stress and competing demands. LLM-based interpretation broadly reproduced the high-level subgroup patterns identified through MATA, but outputs were more generalised, less conceptually differentiated, and showed greater thematic overlap. Bias analysis showed systematic shifts in vocabulary, sentiment, and emotional tone when demographic labels were swapped, suggesting a risk of representational bias.

**Conclusions:** MATA identified meaningful socio-demographic differences while retaining interpretative depth at scale. LLM-based topic interpretation showed utility for rapid thematic summarisation, but produced less conceptually differentiated outputs and was sensitive to demographic framing. The analysis also identified “LLM speak”, where outputs appeared coherent but relied on abstract, generalised, and overlapping interpretations. Human oversight, structured qualitative appraisal, and explicit bias evaluation are necessary when using LLMs to analyse socially stratified free-text health data.

**Author summary:** This study explores how Artificial Intelligence can help researchers analyse very large collections of free-text health experiences without losing important human meaning. We analysed responses from more than 2,000 adults in the United Kingdom who described how the COVID-19 pandemic affected their wellbeing, routines, relationships, and daily lives over time.

We applied a method called Machine-Assisted Topic Analysis, which combines computational topic modelling with human qualitative interpretation, to identify differences across gender, age, and socioeconomic groups. We then compared these findings with analyses generated using a large language model.

Both approaches identified broad patterns in the data, including disruption, uncertainty, adaptation, and the importance of work, family, and health. However, the large language model produced more generalised and overlapping interpretations, whereas the human-led approach retained greater nuance and clearer distinctions between social groups. We also found that changing demographic labels, such as “men” or “women”, systematically altered the language and emotional tone generated by the model, suggesting a risk of representational bias.

Our findings suggest that Artificial Intelligence may be useful for rapid summarisation of large-scale health experience data, but that human oversight remains necessary to preserve nuance and support fair interpretation of socially diverse experiences. As large language models become more widely used in health research and service evaluation, careful evaluation of potential bias will be important to avoid reproducing overly simplified or stereotyped accounts of different social groups.

## Introduction

Free-text data generated in healthcare and public health provide contextualised insights into lived experiences that structured measures often fail to capture. Such data can reveal nuanced concerns and variation across socio-demographic groups, offering insights for equitable healthcare delivery and policy development [1,2]. However, the growing volume of free-text data in digitalised health systems poses significant analytical challenges. Manual qualitative approaches offer interpretive depth but do not scale to large corpora, while automated methods can obscure or distort subgroup variations (especially when such groups are underrepresented) or introduce bias [1].

These challenges become particularly consequential during public health emergencies. During the COVID-19 pandemic, timely population-level understanding was essential to respond to rapidly evolving needs and to ensure interventions addressed socio-demographic disparities. Systematic differences in mood, wellbeing and related stressors, and health outcomes were observed across groups, including variation by age, socioeconomic status, and gender [3–5]. These findings highlight the importance of nuanced, population-level analysis of health-related experiences across socio-demographic groups using large-scale free-text data. However, no scalable and interpretable approach of free-text population experience data-to-public health actionable insights was operationalised in time during the pandemic, and some relevant methods were published too late to inform real-time improvements [6].

Although computational methods have increasingly been applied to free-text data [6–10], existing work does not systematically integrate socio-demographic analysis with interpretable qualitative outputs. A handful of approaches explicitly incorporated socio-demographic information into computational analyses. For example, Shi et al. [11] proposed a graph-based pipeline integrating demographic attributes with unstructured population data. Lin et al. [12] applied demographic inference and post-stratification methods to adjust for biases in Covid-19 related Twitter data and examine demographic disparities in public sentiment. Park et al. [13] integrated demographic metadata with sentiment analysis of online psychiatrist reviews to examine how physician characteristics were associated with patient satisfaction.

Identifying socio-demographic variation in large-scale free-text data is complex because analytic methods may obscure the experiences of under-represented groups, and introduce or amplify bias in socially stratified datasets. The observed differences can depend on the analytic approach applied [14,15], with empirical studies showing that outputs and classifications vary systematically across socio-demographic groups [16–19]. This raises the need for systematic evaluation of analytic approaches when examining socio-demographic variation.

Large language models (LLMs) introduce additional complexity in this regard. Unlike probabilistic topic models, LLMs process text in a context-sensitive manner and can support tasks such as thematic summarisation and code generation [20,21]. However, because model outputs are shaped by patterns in training data, LLM-generated interpretations may reproduce prevailing social and linguistic hierarchies, overrepresenting majority perspectives and attenuating minority experiences [22]. Systematic demographic bias has been documented across state-of-the-art models, with coding errors and output quality varying by respondent characteristics such as gender and education [23]. Further challenges include limited transparency, potential oversimplification of complex qualitative material, and sensitivity to contextual framing, as well as hallucination and topic granularity issues inherent to LLM-based qualitative analysis [24,25].

Responsible deployment of LLMs therefore requires structured human oversight, particularly when socio-demographic variation is central to the research question [20,26].While some scholars have argued for rejecting generative AI in interpretive qualitative research altogether [27],a more productive methodological approach is to test these concerns empirically and develop standards for the use of LLMs in lived-experience free-text analysis. *This study takes this evaluative approach.* This study takes this evaluative approach. Responsible deployment therefore remains contingent on empirical evaluation of whether LLM-based approaches can support valid and unbiased qualitative interpretation, particularly across socio-demographic groups [28,29].

We build on prior work developing analytic approaches that combine computational scalability with interpretive depth and structured oversight, while maintaining transparency and socio-demographic sensitivity. Machine-Assisted Topic Analysis (MATA) integrates computational methods with interpretive human analysis [30,31]. Using AI-based topic modelling, it identifies patterns within large volumes of textual data, which qualitative researchers subsequently interpret and contextualise into meaningful themes [6,30,31]. Previous studies have applied MATA to analyse large-scale free-text data, including survey responses, service feedback, and social media content. Evidence suggests that MATA can improve analytic efficiency while maintaining the interpretive depth of qualitative analysis, particularly in contexts requiring timely analysis on large scale experience data [6,31]. In parallel, the GRACE framework (Grounded Review and Assessment of Computational Evidence) was developed to enable structured evaluation of machine-assisted outputs for qualitative research [32].

The aim of this study was to examine how socio-demographic differences in mood and wellbeing are manifested within a large-scale free-text dataset, and to evaluate how different AI-assisted analytic approaches identify and potentially introduce bias. The study had three primary objectives:

1. To identify potential socio-demographic differences in mood and wellbeing within a large-scale free-text dataset using Machine-Assisted Topic Analysis (MATA), a collaborative AI-human method
2. To systematic compare analytic outputs of MATA (machine learning model +human qualitative analysis) against LLM qualitative analysis (machine learning model + LLM analysis) using GRACE evaluation indicators: interpretability, actionability nuance, redundancy
3. To examine the potential for bias in LLM-based analysis

## Methods

### Dataset: COVID-19 well-being tracker (2,000 free-text responses)

This study draws on free-text data collected from the COVID-19 Wellbeing Tracker Study, a longitudinal survey conducted on a cohort of UK residents aged >= 18 years with access to a smartphone recruited in April 2020 [33,34]. Recruitment was purposive, targeting individuals at higher risk of adverse health outcomes, including those with chronic physical conditions, residents of high-deprivation areas, and individuals with self-reported mental health concerns. Participants enrolled online and completed follow-up surveys at 3, 6, 12, and 24 months post-baseline. Each follow-up included a free-text item: “Please tell us what factors have influenced your health behaviours, mood, and sense of wellbeing over the last [3/12] months.” The aggregated responses to this question constitute the dataset for topic modelling. The raw dataset contained 4,176 entries which was then filtered to 2,177 entries following standard preprocessing and quality assurance procedures [31].

### Experimental Setup

All experiments were conducted using Python 3.11, with Pandas for data processing, NLTK for text preprocessing, and scikit-learn for machine learning tasks.

### Objective 1 - MATA

#### Data preprocessing

##### Text Preprocessing

Responses were lowercased, and non-alphabetic/non-ASCII characters removed. Text was tokenised by whitespace, with standard English stop words (NLTK corpus) and domain-specific terms removed, including ‘covid’ (ubiquitous and uninformative given the study context) and contraction fragments (e.g., ‘im’, ‘ive’, ‘fa’) resulting from non-alphabetic character removal. Remaining tokens were lemmatised using the NLTK WordNet lemmatiser. Empty documents were excluded.

##### Document-Term Representation

Preprocessed text was vectorised using scikit-learn’s CountVectorizer, filtering terms appearing in over 95% of documents (uninformative due to ubiquity) or fewer than 2 documents (too rare to contribute to generalisable topic patterns), producing a document-term matrix for input to the LDA model.

- **Stratification**: **Gender (male, female):** Recorded numerically (1 = male, 2 = female). Analyses were restricted to these two categories to ensure adequate subgroup sizes.
- **Socioeconomic status (SES):** SES was derived from Index of Multiple Deprivation (IMD) deciles linked to residential postcodes. IMD is a composite measure of area-level deprivation (income, employment, education, housing, health) with data obtained from IMD indexes for England (2019), Northern Ireland (2017), Scotland (2020), and Wales (2019). Participants were dichotomised into higher deprivation (deciles 1–6) and lower deprivation (deciles 7–10). Thei division approximates a contrast between relatively less deprived areas and the remainder of the socioeconomic distribution which is otherwise less pronounced. Cases with missing IMD were excluded from SES-stratified analyses.
- **Age group (18–49, 50+):** Participants were categorised using predefined age bands. As eligibility was restricted to adults, the under-18 category was negligible and excluded.

### Latent Dirichlet Allocation

We adopted Latent Dirichlet Allocation (LDA) [35], which represents each document as a probabilistic mixture of latent topics, allowing transparent inspection of topic–word and document–topic distributions. Given the moderate corpus size (2,177 responses) and the study’s focus on explainable insights, LDA provided an appropriate balance between rigour and interpretive clarity.

Separate LDA models were trained per subgroup to capture group-specific thematic structures, as topics are not assumed to be shared across demographic groups.

For each topic, the ten highest-weighted terms were extracted to characterise thematic content, and the 20 responses with the highest topic probability were identified as representative texts.

The top five representative responses per topic were passed through a pre-trained BART-large-CNN abstractive summarisation model [36], generating individual summaries that were concatenated to produce a combined narrative per topic.

### Qualitative analysis methods

To enable expert interpretation of the topics, we conducted a human-only qualitative analysis phase. We extracted the topic modelling outputs from the model and presented them to two qualitative researchers with extensive experience in qualitative health research, traditional qualitative methods, and large-scale free-text data analysis (EB, EW).

Researchers were blinded to socio-demographic subgroup identifiers. Datasets were anonymised and labelled neutrally in Excel to minimise interpretive bias. As per MATA approach [30] for each subgroup (gender, SES, and age), 20 representative responses per topic were selected based on the highest document–topic probabilities.

EB and EW independently reviewed extracts, writing brief descriptive notes and inductively generating initial codes based on recurring meanings within each topic and subgroup. Throughout the process, researchers maintained analytic memos to document interpretive decisions, reflections on topic clarity, perceived difficulty of interpretation and any other thoughts about the process.

Researchers then met to compare interpretations, discuss discrepancies. Finally, researchers iteratively refined labels to reflect analytically meaningful topics. Researchers reviewed similarities and differences of individual topics within each socio-demographic group (gender, SES, and age), clustering topics across subgroups for shared sematic meaning.

### Objective 2 - Large Language Model as a Qualitative Researcher

We compared Machine-Assisted Topic Analysis (MATA) with topic modelling combined with LLM-based topic interpretation. We developed a structured prompt in consultation with qualitative researchers to guide the LLM analysis of topic model outputs.

For each topic within each demographic subgroup, the 20 representative responses with the highest topic probability were passed to GPT-4o-mini (temp=1.0, top_p=1.0; selected for its cost-efficiency and scalability) via the Azure OpenAI service. The model was prompted to act as a qualitative researcher and was given the research context: identifying what influenced participants’ mood, behaviour, and well-being during COVID-19. For each topic, it was asked to provide an overarching theme name, a short description of what characterised the topic, and one to three illustrative excerpts drawn from the representative responses (Appendix 1 for prompts).

The LDA topic labels were agreed by two researchers using the MATA process and the LLM topic labels were generated by the model. After reviewing the analyses, researchers grouped the topics within into clusters for shared sematic meaning. Some of the clusters bridged subgroups, whereas some clusters or standalone topics were only within one subgroup. This process was completed for each model to evaluate similarities and differences within groups and between MATA and LLM “interpretation”.

Evaluation of the outputs using GRACE - Grounded Review and Assessment of Computational Evidence

During the analytical process, two qualitative researchers independently evaluated the outputs of: 1) the 6 LDA models, and 2) the 6 LLM “interpretations”, using the standard GRACE procedure (see [37] for details).

GRACE is a systematically developed and evaluated framework for assessing the quality of AI-generated outputs from textual data analysis. It comprises four indicators: interpretability, actionability, nuance, and redundancy. The framework combines a structured scoring system with qualitative interrogation through free-text justifications.

The researchers (EW, EB, PB) then met to discuss the evaluations and reach consensus. They reviewed the scores, notes, and meeting transcripts, and completed the GRACE evaluation collaboratively.

### Objective 3 - Large Language Model Bias Analysis

To assess potential bias in LLM qualitative outputs across sociodemographic groups, we tested whether LLM-generated themes varied as a function of the stated demographic label in the prompt rather than the underlying data.

Bias in large language models (LLMs) is typically detected via quantitative benchmarking, controlled prompting, or embeddings auditing [38]. Standard approaches involve evaluating model outputs on curated bias benchmarks, such as stereotype completion tasks or fairness-oriented QA datasets, which measure systematic differences in responses across demographic groups [39,40].

Counterfactual testing is widely used, where sensitive attributes are varied in otherwise identical prompts to assess whether outputs change inappropriately [41,42]. In addition, embedding-based analyses can reveal latent biases encoded in the model’s internal representations [43,44]. We focus on the accessible and widely accepted counterfactual evaluation.

For each stratified LDA output, the 20 representative responses and topic keywords from a given subgroup were provided to GPT-4o-mini (temp=1.0, top_p=1.0), instructed to conduct thematic analysis with the data labelled as that subgroup (e.g., “female participants”). The analysis was repeated with an identical prompt but the demographic label swapped to its counterpart (e.g., “male participants”; counterfactual condition). This was carried out for both subgroups within each demographic dimension: gender (male vs female), SES (IMD deciles 1–6 vs 7–10), and age (18–49 vs 50+).

Each subgroup contributed five LDA topics from a separate model. For each topic, the LLM generated 20 outputs under the correct label (true condition) and 20 under the swapped label (counterfactual condition), yielding 40 outputs per topic, 200 per subgroup, and 400 per dimension. A non-zero temperature introduced stochastic variation across iterations. Because both conditions share identical data and differ only in the demographic label, systematic differences in vocabulary or framing can be attributed to the label itself.

To quantify narrative shift between conditions, we conducted a lexical analysis measuring the degree to which vocabulary and word usage patterns change when the label is swapped. A biased model would exhibit significant lexical shifts when describing the same data for different demographic groups, whereas an equitable model would maintain a stable core vocabulary.

All text was preprocessed using NLTK: tokenised to lowercase, with stop words and non-alphanumeric tokens removed. We employed two metrics to capture lexical similarity between aggregated token pools from all true-condition versus all counterfactual-condition texts for the same subgroup.

– Jaccard index: This metric assesses the overlap of the unique vocabulary between two sets. It is calculated as the size of the intersection of the two sets of unique words divided by the size of their union.

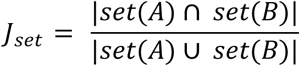

– Cosine similarity was computed over term-frequency vectors, where each list was represented as a vector of word counts across the combined vocabulary of both lists. The cosine of the angle between these vectors quantifies the degree of lexical overlap, with values closer to 1 indicating more similar word usage patterns.

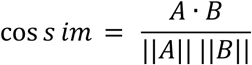

We further implemented a bootstrap resampling strategy for each subgroup and topic:

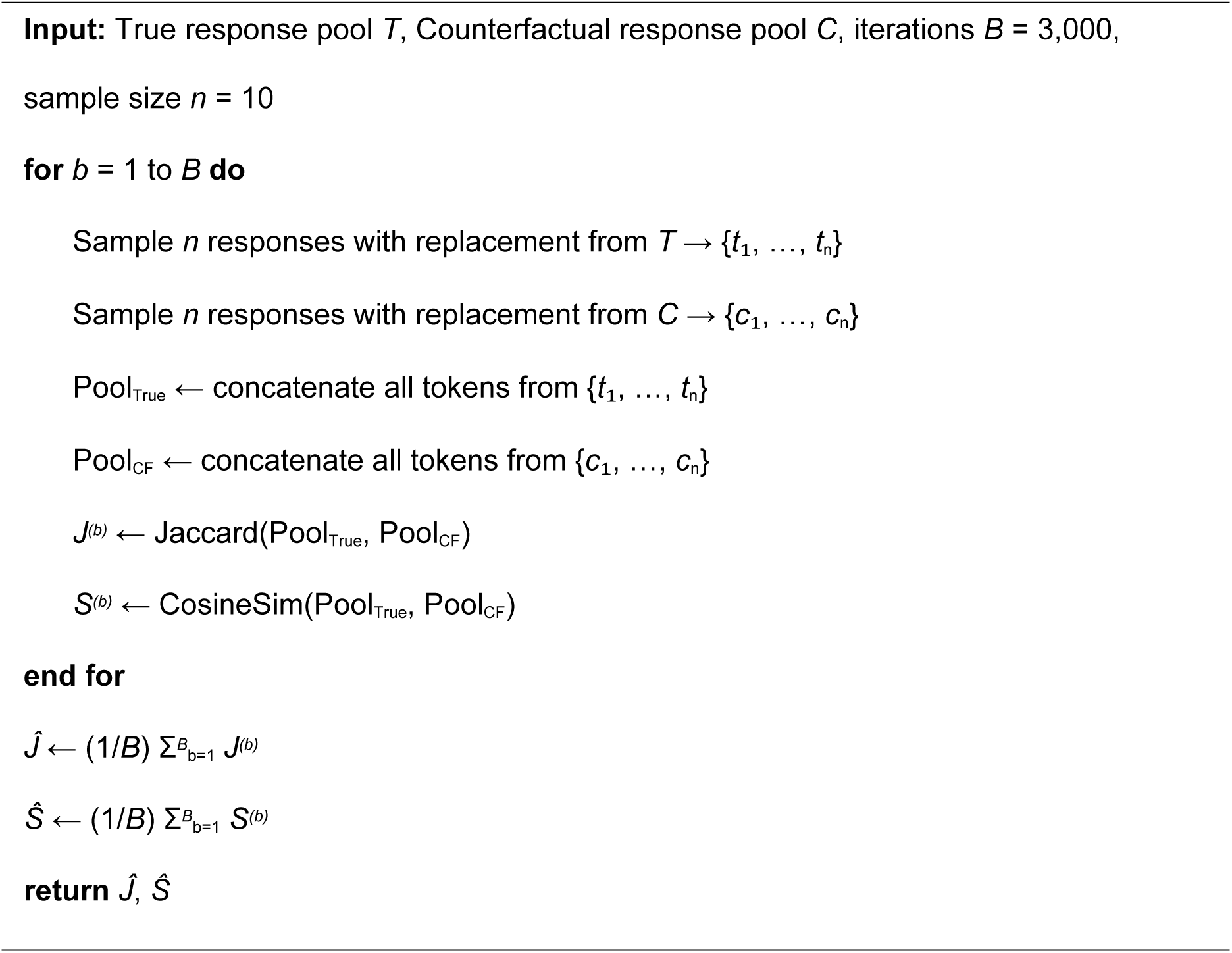

We implemented a bootstrap resampling strategy to estimate lexical similarity between true and counterfactual conditions for each subgroup–topic pair. The procedure is detailed in Algorithm 1. For each pair, B = 3,000 bootstrap iterations were used to ensure stable similarity estimates, following standard recommendations for confidence interval estimation [45]. At each iteration, 10 responses were sampled with replacement from each condition’s pool, tokens were concatenated, and Jaccard index and cosine similarity were computed between the resulting pools. The mean across all iterations yields the Observed Similarity (Obs_Jaccard, Obs_Cosine) for that subgroup–topic pair. This procedure was applied independently across all 30 subgroup–topic pairs (3 demographic dimensions × 2 subgroups × 5 topics).

To contextualise the Observed Similarity scores, we computed a Baseline Similarity capturing the inherent lexical variability within a subgroup’s own true-condition responses. The same bootstrap procedure (Algorithm 1) was applied, but both pools were drawn from the same subgroup’s true-condition responses rather than comparing true to counterfactual pools. Specifically, for each iteration, two independent samples of 10 responses were drawn with replacement from the true pool (Pool_True_A and Pool_True_B), and Jaccard index and cosine similarity were calculated between them. The mean across B = 3,000 iterations forms the Baseline Similarity (Base_Jaccard, Base_Cosine) for that subgroup.

We employed VADER (Valence Aware Dictionary and Sentiment Reasoner [46]) to assess overall affective tone. VADER compound scores were computed at sentence level within each output and averaged to obtain a single sentiment value, preserving VADER’s sensitivity to negation and intensifiers. A sentiment delta was computed for each iteration pair:

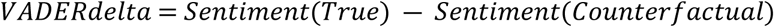

To capture more granular emotional dimensions beyond unidimensional valence – since overall sentiment may remain stable while the type of affect shifts (e.g., fear-framed versus anger-framed negativity) – we utilised the NRC Emotion Lexicon [47], containing eight basic emotions: anger, anticipation, disgust, fear, joy, sadness, surprise, and trust. For each iteration pair, we identified the shared vocabulary between true and counterfactual texts and isolated the differential tokens – words exclusive to each condition, with repetitions preserved. This focuses the analysis on where the LLM’s language diverges between conditions. Emotion densities were calculated over these differential token pools:

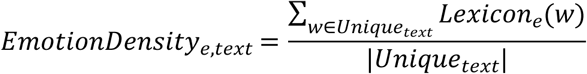

Here, emotion density represents the proportion of condition-exclusive words carrying a given emotion, where higher values indicate a greater concentration of that emotion in the LLM’s distinctive vocabulary for that condition.

Emotion delta scores were then computed for each of the eight dimensions to quantify the directional shift in each emotion’s prevalence between conditions:

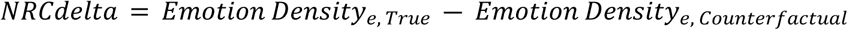

## Results

We start by presenting the results of the differences observed between the gender, SES, and Age for Objective 1 for MATA (6 LDA models + qualitative analysis), and Objective 2 same LDA models, followed by LLM-generated “interpretation”. We then compare the thematic similarities and differences identified through MATA and LLM interpretation, summarised in the table below.

### Objective 1 MATA: 6 LDA models + qualitative analysis

Below are agreed topic labels agreed by two qualitative researchers using the MATA process by socio-demographic stratification (blinded qualitative analysis).

### Differences identified through MATA analysis

#### Gender

Connection, isolation and health was prominent in the female-stratified analysis, with focus on physical and mental health difficulties, caregiving, isolation, and concern for vulnerable family members. Accounts were more introspective and emotional, with explicit anxiety related to health, COVID risk, family, and the future.

In contrast, male-stratified topics emphasised work stress, disrupted routines, and changing relationships shaping mood and sense of control. Coping centred on establishing routines and habits, with narrative weight on managing external pressures, supported by relationships.

#### SES

Lower SES topics emphasised barriers and facilitators to wellbeing, with focus on financial worries, unstable work, health, and immediate practical constraints.

Higher SES topics emphasised loss and connection, including bereavement, shielding, and social restrictions linked to low mood, with adaptations to maintain connection (e.g. online communication). Narratives reflected meaning-making and expectations of autonomy.

#### Age

In the 50+ group, topics focused on maintaining wellbeing through routines and health behaviours, with health concerns prominent. Relationships featured through concern for others, lack of support, and family separation, contributing to isolation and anxiety.

In the 18–49 group, topics focused on external demands and mental health. Pressures across work, home, caregiving, and health led to exhaustion and stress, with substantial impact on mental health and, for some, reduced motivation for health behaviours.

### Objective 2 LLM analysis - 6 LDA models + LLM “interpretation”

Differences generated by LLM “interpretation” summarised by qualitative researcher Gender

Male topics emphasised resilience and coping through routines, health behaviours, and support networks, primarily in response to work and health pressures. Female topics focused on connection, isolation, and health, highlighting disrupted relationships, loneliness, and emotional strain. Additional emphasis was placed on physical and mental health challenges affecting both self and others.

#### SES

Lower SES topics emphasised maintaining social connections as protective for wellbeing.

Higher SES topics incorporated broader societal influences (e.g. government response, media, future uncertainty), linking mental health to both personal and structural factors.

#### Age

18–49 topics centred on balancing work, caregiving, and personal wellbeing, with cumulative pressures leading to stress and fatigue. Adaptation involved routines and maintaining relationships.

50+ topics focused on relationships and health, including family dynamics, isolation, and health concerns. Adaptation centred on maintaining connections and managing health.

#### Comparison of thematic similarities and differences between MATA and LLM

For both LDA and LLM analysis, women’s accounts emphasised more relational, health and emotional impacts, while men emphasised routine-based coping, whilst both genders shared themes of pandemic related demands. LDA analysis revealed a female topic related to pandemic restrictions; this was not an identified topic in the LLM analysis. Table 1 shows the topic labels for both the LDA and LLM analysis.

**Table 1:** Topic labels for MATA (LDA + human qualitative analysis) and LDA + LLM “interpretation” showing similarities and differences within the sociodemographic variables and between the MATA and LLM model analysis.

|  | LDA topic labels |  | LLM topic labels |  |
| --- | --- | --- | --- | --- |
| Gender | Men | Women | Men | Women |
| Similarities | **Demands cluster:**<br>External pressures and personal resilience during: health, work, political and social influences<br>Lockdown disruption and demands: attempting to balance life's pressures | Lockdown demands: Maintaining stability, well-being, and a sense of purpose | **Demands cluster:**<br>Balancing Mental Health and Well-being Amidst Stressors during COVID-19<br>Balancing Responsibilities and Personal Well-being during COVID-19 | Coping Amidst Uncertainty |
| Differences | **Health and mental health**<br>facilitators and constraints and the interactions between them | **Pandemic restrictions,** risk and resilience |  | **Health cluster:**<br>Health Challenges and Mental Well-being During COVID-19<br>Health, Worry, and the Impact of Isolation |
|  | **Routines and habits cluster:**<br>Striving for routines and ability to regain control amid lockdown restrictions<br>Maintaining healthy habits in face of change | **Connection, isolation and health cluster:**<br>Impacts, influences and effects and health and wellbeing of them and their family<br>Anxieties and enduring isolation and health struggles<br>Reflections on connection and adaption during the pandemic | **Resilience and coping cluster:**<br>Personal Resilience and Coping Mechanisms During COVID-19<br>Resilience and Adaptation During Adversity<br>Health, Well-Being, and Social Connections | **Relationships and connection cluster:**<br>Connection and Coping Amidst Chaos<br>The Impact of Isolation and New Routines on Well-Being |
| SES | Lower SES | Higher SES | Lower SES | Higher SES |
| Similarities | **Change, adaption and reflection cluster:**<br>Level of adaption to significant life changes<br>Reflections on the impact of lockdown | Restricted autonomy and ability to adapt | **Family dynamics and relationships cluster:**<br>Struggles with Health and Family Dynamics During COVID-19 | Navigating Health and Well-Being Amidst Work and Lockdown Challenges<br>Family Struggles and Social Isolation |
|  | **Demands, loss of control, and mental health cluster:**<br>Mental health impact and influences | Mental health impacts from loss of control | **Demands and pressures cluster:**<br>Adjustments and Challenges in Daily Life During COVID-19<br>Balancing Home, Work, and Family Responsibilities | Health on Emotional Well-being During COVID-19<br>Balance of Work and Well-being at Home |
|  | Emotional and practical demands and loss of control during the pandemic |  | Balancing Home, Work, and Family Responsibilities |  |
| Differences | **Loss and connection** during the pandemic | **Interactions between factors** that help and hinder individuals<br>**Making the best of it:** resilience and navigating uncertainty | **Connecting Through Relationships** During Isolation | Complex interplay of **Personal Circumstances and Wider Societal Factors** on Mental Health during COVID-19 |
| Age | 18-49 years | 50+ years | 18-49 years | 50+ years |
| Similarities | **Adaption and coping cluster:**<br>Disruption to personal autonomy: reflections and re-evaluation<br>Adjustment during times of uncertainty and change | Appraisal of coping with the restrictions<br>Adaptation and adjustment to restrictions |  |  |
|  | **Purpose and control cluster:**<br>Ability to seek purpose | Maintaining a sense of purpose/routine and coping mechanisms |  |  |
| Differences | **Motivation and coping** with mental health | **Factors impacting health and wellbeing** and interactions between them | **Adaption and coping cluster:**<br>Balancing Personal Well-being and External Pressures During COVID-19<br>Coping and Resilience amidst Challenges<br>Adapting to Change: Balancing Health, Work, and Well-being During COVID-19 | **Health cluster:**<br>Challenges of Home and Health during COVID-19<br>Family Dynamics and Personal Health during COVID-19<br>Resilience and Adaptation Through Family Support and Health Awareness |
|  | **Balancing external pressures** | Professional and personal **connection** and its impact on wellbeing | **Balancing demands:**<br>Balancing Responsibilities and Well-Being During COVID-19<br>Balancing Personal and Professional Well-being in the Context of Isolation and Uncertainty | **Connection and relationships:**<br>Connections and Activities at Home: Adapting to Life During Lockdown<br>Isolation and Family Dynamics During COVID-19 (50+T2) |

Both LDA and LLM analyses identified shared pandemic-related demands across SES groups, but highlighted differences in experience, with more deprived groups emphasising financial and practical constraints and less deprived groups focusing on emotional and existential impacts. Relationships and connection were important across analyses, particularly for more deprived groups. The LDA analysis revealed more lockdown adaption focussed topics compared to LLM.

The LDA analysis identified themes related to adaption and coping across both age groups, whereas this was related to the younger age group only for the LLM analysis. The LDA analysis explicitly linked this theme to lockdown restrictions, whereas it was presented more generally in the LLM analysis. Both models highlighted topics related to health and connection in the older age groups and balancing demands and pressures in the younger age groups. The LDA analysis also identified topics related to purpose and control themes.

#### GRACE evaluation of MATA and LLM outputs

GRACE scores were applied to the final thematic outputs generated by each approach (MATA and LLM), enabling comparison of the interpretive quality of outputs derived from the same dataset, despite differences in analytic process. The components of GRACE (assessed by two researchers) evaluation are summarised below. We start by presenting GRACE scoring followed by qualitative evaluation.

MATA performed better overall for gender and SES, particularly on interpretability and nuance (Table 2). The strongest performance was observed for the gender-stratified MATA output. The age group analysis was scored the same overall for both models, with LDA performing better on nuance than the LLM, but worse on interpretability than the LLM and the other LDA group analyses. This was reflected in the qualitative evaluation with researchers noting multiple difficulties with interpretation for age. Overall, findings were comparable, with LLM outputs offering broader interpretive insights and LDA-derived outputs providing more granular distinctions across groups.

**Table 2.**
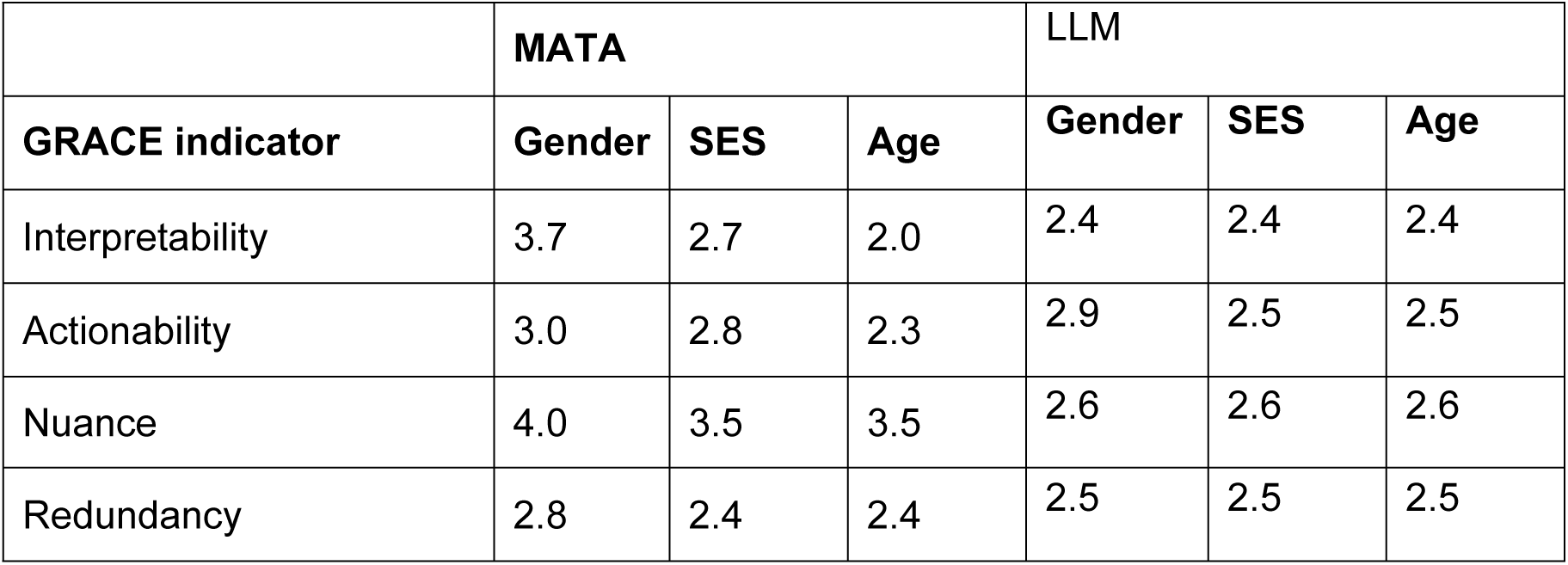
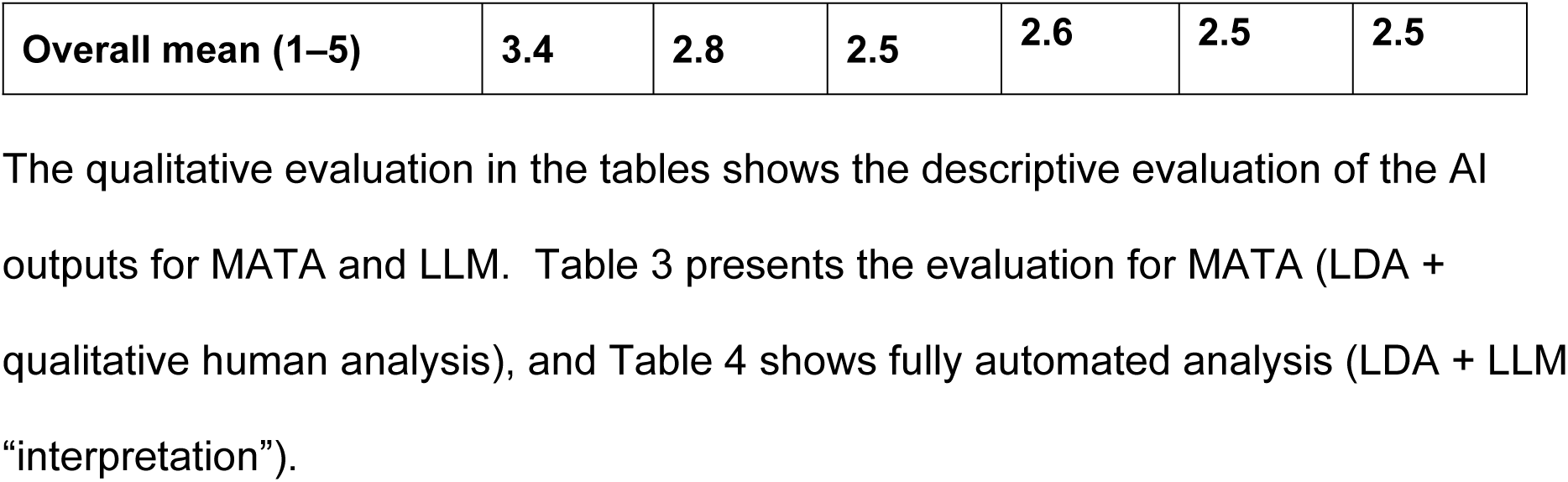
GRACE scores for LDA output LLM outputs by stratification.

The qualitative evaluation in the tables shows the descriptive evaluation of the AI outputs for MATA and LLM. Table 3 presents the evaluation for MATA (LDA + qualitative human analysis), and Table 4 shows fully automated analysis (LDA + LLM “interpretation”).

**Table 3:** Qualitative evaluation of the LDA models’ performance using GRACE.

| **GRACE domain** | **LDA–Gender** | **LDA–SES** | **LDA–Age** |
| --- | --- | --- | --- |
| **Interpretability** | Quotes generally aided interpretation, although they were often long and contained multiple ideas. Confidence improved after the consensus discussion. | Interpretation required more effort than for gender. Quotes were often lengthy and included several subthemes, which reduced clarity and made rapid interpretation more difficult. | Topics were longer and more complex, making interpretation slower and sometimes difficult. Quotes frequently contained multiple ideas, requiring careful reading and discussion to reach consistent interpretations. |
| **Actionability** | Topics provided some conceptual insights related to the research question but remained broad. Findings would need triangulation with other research to be useful for policy and were limited on their own. | Findings highlighted some differences across SES groups, though these were generally subtle and descriptive. Additional interpretation or triangulation would likely be needed to support policy relevance. | Outputs provided some descriptive insights but were less directly aligned with policy-oriented questions. The age split appeared less informative for practical decision-making. |
| **Nuance** | Topics captured nuanced perspectives and some distinctive viewpoints in the data, contributing to their interpretive value. | Topics reflected different perspectives across SES groups, although distinctions between themes were sometimes less clear. | Topics also reflected nuanced content, although the complexity of the themes sometimes made these distinctions harder to identify quickly. |
| **Redundancy** | Some semantic overlap between topics blurred boundaries | Overlap between topics was more apparent, with some | Redundancy was also present in the age-stratified topics, with |
|  | between themes, although the main thematic areas remained broadly distinguishable. | themes appearing conceptually similar within and across SES categories. | recurring themes reducing overall distinctiveness. |

**Table 4:** Qualitative evaluation of the fully automated AI analysis, using GRACE.

| Domain | LLM – Gender | LLM – SES | LLM – Age |
| --- | --- | --- | --- |
| **Interpretability** | Inconsistent topic structure, shallow descriptions, limited number of quotes, constraining verification; Topic labels often broad or loosely aligned with descriptions ; Summary sentences rely on vague phrasing (e.g. “**rich tapestry**”, “**multifaceted nature**”) ; Some **quotes** misaligned or include unaddressed subthemes ; Evidence of truncation raises concerns about excerpt selection ; Overlapping topics reduce clarity and separability | Inconsistent presentation, sparse quotes, broad or weakly specified descriptions ; Topics combine concepts without explanation; unclear boundaries ; Interchangeable use of mental health, physical health, wellbeing reduces conceptual precision ; Limited transparency in quote selection ; Topic overlap reduces analytic clarity | Inconsistent structure, shallow thematic descriptions, limited quotes ; Labels and summaries often broad; mismatch between topic names and content ; Use of “**double concept**” (e.g. health + connection) merges distinct ideas without explanation ; Reduces clarity and weakens boundaries between themes ; Topic overlap further limits interpretability |
| **Actionability** | Broad alignment with gender differences (male: routine/adaptation; female: communication/connection/health/anxiety) ; Descriptions too general; limited analytical usefulness and lockdown-specific insight ; May serve as **headline findings**; potential for gender-sensitive wellbeing support ; Lacks depth for meaningful insight or stronger practical application ; Likely reflects expression differences rather than mechanisms ; Quote truncation and underdeveloped descriptions reduce confidence | Weakest actionability ; Some actionable specifics (e.g. working from home), but lacks depth for intervention design ; Underdeveloped descriptions; weak practical utility ; Differences (e.g. social vs broader concerns) insufficient for decision-making ; May reflect grouping approach rather than substantive insight ; Remains at surface-level; limited theoretical and practical value | Relevant age differences (18–49: disruption; 50+: health/connection), similar to LDA ; Themes require restructuring; descriptions underdeveloped ; Limited analytical depth; largely descriptive ; Some potential for intervention insight, but remains surface-level ; Likely reflects expression differences rather than mechanisms ; Truncation reduces confidence for decision-making |
| **Nuance** | Some variation in strength, but overall limited detail and reliance on abstract language ; Themes remain surface-level, insufficiently grounded in examples ; Frequent generalisation; only occasional detailed description ; Binary framing (coping vs struggling) fails to capture co-existing experiences ; Example: coexistence of solace and disconnection not adequately represented; Some nuance present in quotes, but not reflected in descriptions ; Positive and negative experiences presented, often polarised rather than integrated ; No overt stereotyping; some gendered patterning (routines vs relationships), but unclear if data-driven ; Limited quotes constrain evaluation | Weakest nuance ; Highly general descriptions, especially around “demands” ; Strong generalisation and flattening across analysis ; Limited retention of divergent or less dominant experiences ; No clear stereotyping, but limited transparency restricts judgement ; Overall too broad to capture layered experiences | Similarly limited nuance; themes remain surface-level ; Broad summaries flatten experience ; **“Double concept”** themes further obscure distinctions ; Some nuanced insights in quotes, not carried into descriptions ; No overt stereotyping; limited depth and transparency restrict assessment |
| **Redundancy** | Redundancy evident across topics ; Interactions between experiences implied, not clearly articulated ; Significant overlap within gender topics ; Some topics overly broad, repeating concepts ; Imbalance between broad/underdeveloped and more specific topics ; Granularity suboptimal, though **headline findings** remain identifiable | Redundancy present ; Topic overlap persists; repeated concepts across themes ; Some interactions visible in quotes, not reflected in descriptions ; Poor granularity ; Overlapping “demands” topics weaken delineation and distinctiveness | Similar redundancy pattern ; Interactions implied but not clearly described ; Overlap persists; repetition driven by broad thematic framing ; Concepts recur without clear differentiation ; Imbalance in topic specificity; granularity not well calibrated |

#### Retrieval of representative example excerpts

Manual verification of all 90 LLM-generated example excerpts suggested that the excerpts largely remained aligned with the source text. Modifications were typically minor in nature. The most prevalent feature was unindicated truncation, observed in 66.7% of excerpts. A small number of excerpts appeared to combine content from multiple sources. A detailed breakdown of these modification types is provided in Appendix 2.

### Objective 3: LLM Bias Lexical Similarity Analysis

Table 5 presents the bootstrap lexical similarity estimates. Across all three demographic dimensions, Observed Jaccard similarity (0.386–0.395) was substantially lower than Baseline (0.559–0.563), with non-overlapping 95% confidence intervals, indicating that the label swap consistently altered which words the LLM used. Gender showed the largest gap (0.386 vs 0.561), followed by Age (0.388 vs 0.559) and SES (0.395 vs 0.563). Cosine similarity was also lower in the Observed condition (0.876–0.903) than Baseline (0.929–0.940), though the CIs overlapped. Together, these results suggest that swapping the demographic label changes the LLM’s vocabulary (lower Jaccard), but the relative frequency with which words are used remains broadly similar (overlapping Cosine CIs).

**Table 5.**
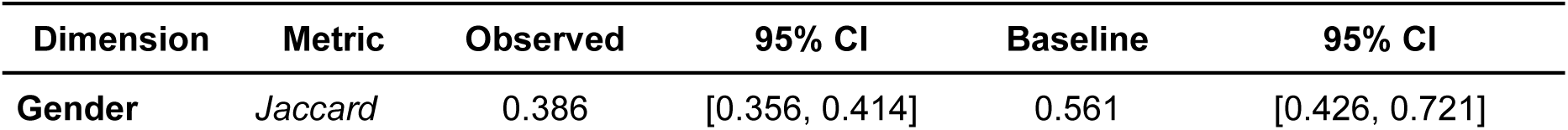
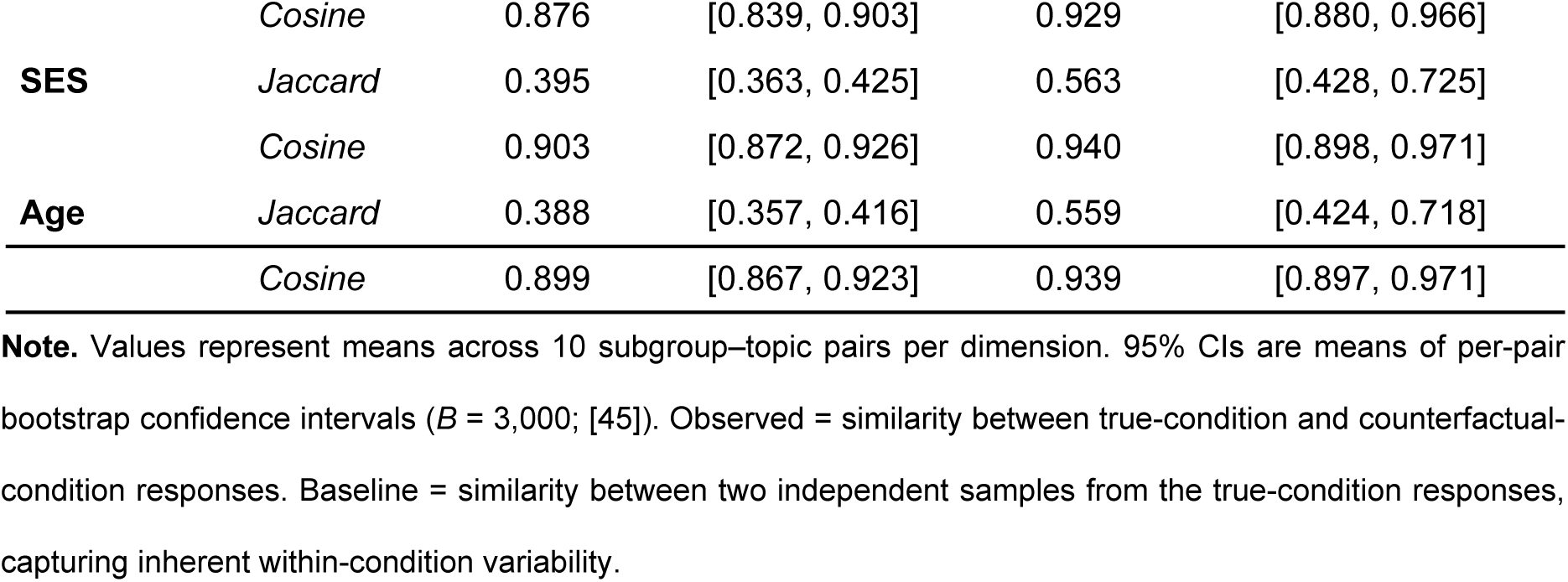
Bootstrap Lexical Similarity: Observed (True vs Counterfactual) and Baseline (True vs True) with 95% Confidence Intervals.

#### LLM Bias VADER Sentiment Shifts

To test whether the sentiment shift for each subgroup was significantly different from zero, we conducted one-sample t-tests on the VADER compound score deltas (True − Counterfactual) for each subgroup. Five of six subgroups showed significant sentiment shifts (Table 6). When the LLM was told the data came from male participants, its output was more negatively toned than when the same data was labelled as female (Δ = −0.042, p = 0.032). Similarly, the lower SES label produced more negative sentiment than the higher SES label (Δ = −0.030, p = 0.043). Conversely, labels for female (Δ = +0.036, p = 0.048), older adults (Δ = +0.050, p = 0.004), and higher SES (Δ = +0.055, p < 0.001) shifted sentiment in a more positive direction. Only the younger age group showed no significant shift (Δ = −0.004, p = 0.802).

**Table 6.** VADER Sentiment Delta (True − Counterfactual) by Subgroup. One-sample t-tests against zero (n = 100 per subgroup).

| Subgroup | Mean $\Delta$ | SD | SEM | t | p |
| --- | --- | --- | --- | --- | --- |
| Gender: Male | -0.042 | 0.191 | 0.019 | -2.180 | 0.032* |
| Gender: Female | +0.036 | 0.181 | 0.018 | 2.006 | 0.048* |
| Lower SES | −0.030 | 0.146 | 0.015 | −2.052 | 0.043* |
| Higher SES | +0.055 | 0.147 | 0.015 | 3.741 | <0.001*** |
| Age 18–49 | −0.004 | 0.164 | 0.016 | −0.252 | 0.802 |
| Age 50+ | +0.050 | 0.171 | 0.017 | 2.942 | 0.004** |

**Note.** $\Delta$ = Mean VADER compound score difference (True – Counterfactual). \* $p < 0.05$ , \*\* $p < 0.01$ , \*\*\* $p < 0.001$ .

Further NRC Emotion Analysis revealed specific lexical choices underlying these sentiment shifts, with significant effects concentrated in Gender and SES (Figure 2). For males (Figure 2a), the true label elicited less joy-related (Δ = −0.007, p = 0.004) and trust-related language (Δ = −0.009, p = 0.004), but more disgust-related language (Δ = +0.003, p = 0.046). For females (Figure 2b), the counterfactual condition elicited more sadness (Δ = −0.007, p = 0.038) and disgust (Δ = −0.004, p = 0.040) compared to the true label. For lower SES (Figure 2c), the true label produced significantly more fear (Δ = +0.009, p = 0.006) and disgust (Δ = +0.004, p = 0.050) language. No significant emotion shifts were observed for Age or higher SES subgroups. Table 7 presents top emotion words, by subgroup.

**Figure 1:**
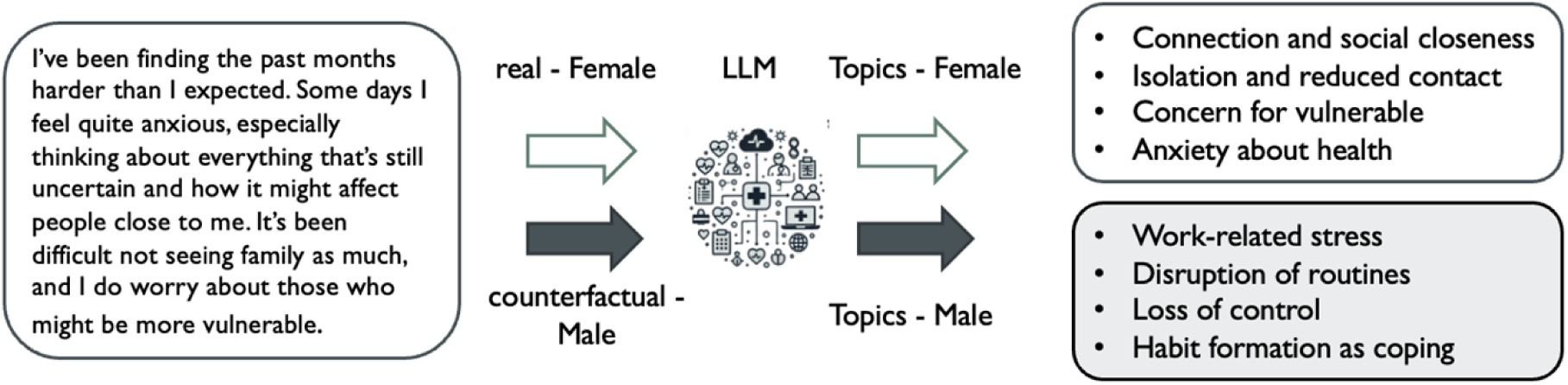
A label-swap crossover design to test demographic framing bias in LLM interpretation. Hypothetical example. Similar experiments are conducted across demographic subgroups.

**Figure 2:**
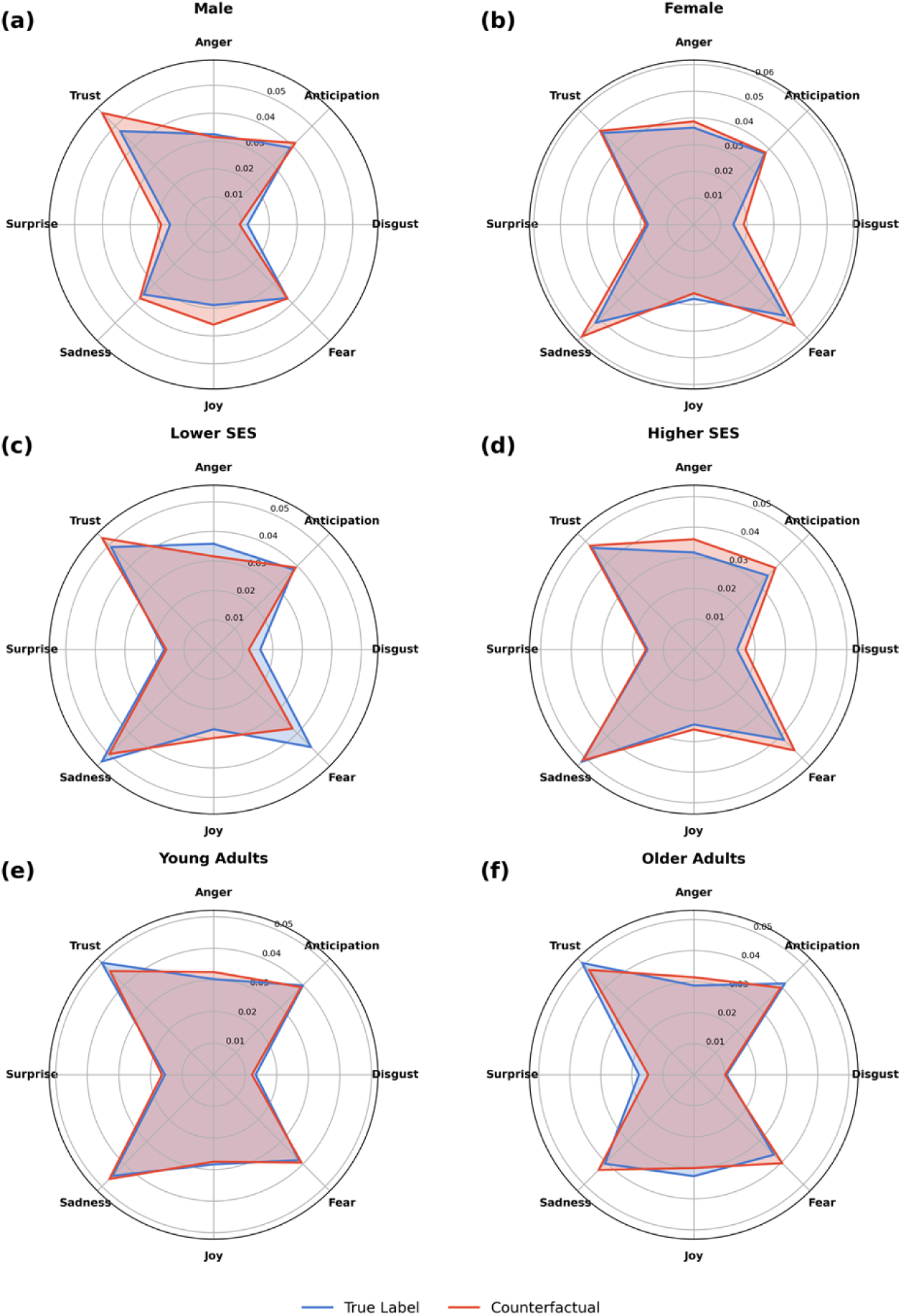
LLM Bias NRC Emotion Analysis

**Table 7:**
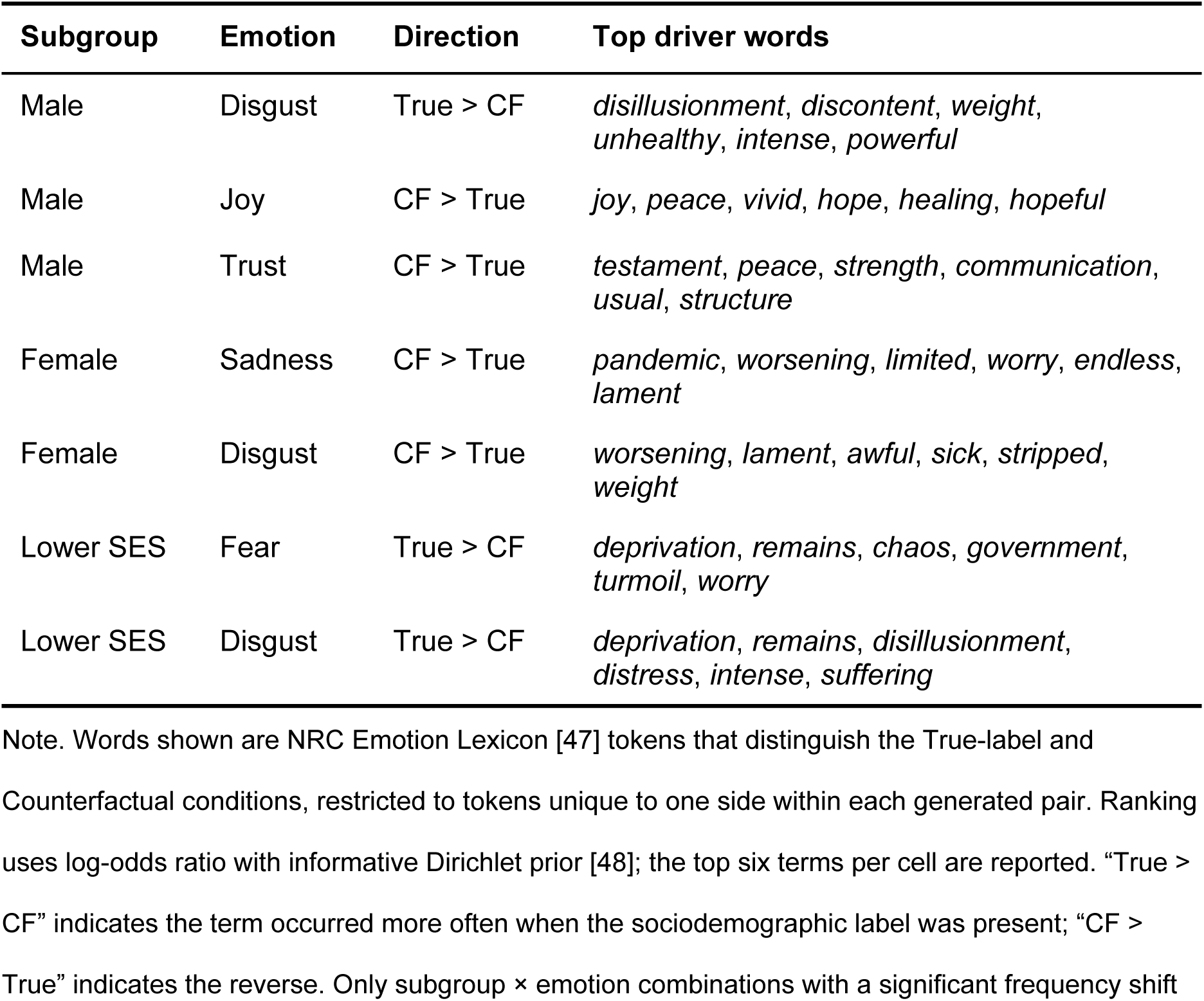
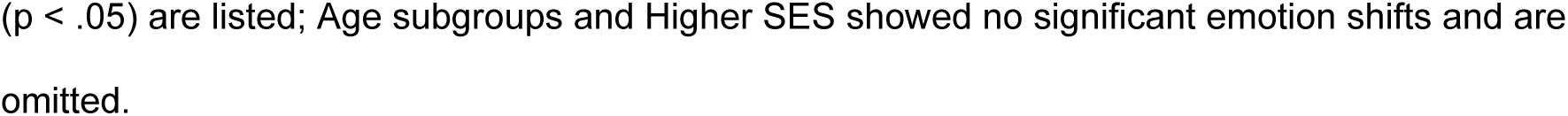
Top NRC emotion words driving significant condition differences, by subgroup.

| Subgroup | Emotion | Direction | Top driver words |
| --- | --- | --- | --- |
| Male | Disgust | True > CF | *disillusionment, discontent, weight, unhealthy, intense, powerful* |
| Male | Joy | CF > True | *joy, peace, vivid, hope, healing, hopeful* |
| Male | Trust | CF > True | *testament, peace, strength, communication, usual, structure* |
| Female | Sadness | CF > True | *pandemic, worsening, limited, worry, endless, lament* |
| Female | Disgust | CF > True | *worsening, lament, awful, sick, stripped, weight* |
| Lower SES | Fear | True > CF | *deprivation, remains, chaos, government, turmoil, worry* |
| Lower SES | Disgust | True > CF | *deprivation, remains, disillusionment, distress, intense, suffering* |

Note. Words shown are NRC Emotion Lexicon [47] tokens that distinguish the True-label and Counterfactual conditions, restricted to tokens unique to one side within each generated pair. Ranking uses log-odds ratio with informative Dirichlet prior [48]; the top six terms per cell are reported. “True > CF” indicates the term occurred more often when the sociodemographic label was present; “CF > True” indicates the reverse. Only subgroup × emotion combinations with a significant frequency shift
( $p < .05$ ) are listed; Age subgroups and Higher SES showed no significant emotion shifts and are omitted.

## Discussion

### A statement of overall findings

This study produced three main findings. First, machine-assisted topic analysis (MATA), combining topic modelling with human qualitative interpretation, identified meaningful socio-demographic thematic differences in pandemic-related mood and wellbeing across gender, socioeconomic groups (SES), and age. Second, when this interpretative stage was replaced with LLM-generated “interpretation”, subgroup differences remained broadly recognisable but became less conceptually differentiated. Third, LLM-generated qualitative analysis was sensitive to demographic cues: the same underlying content produced systematic shifts in vocabulary and emotional tone depending on the demographic label applied, suggesting a risk of representational bias.

Across all three stratifications, the LLM outputs remained broadly plausible, but repeatedly defaulted to high-level categories such as coping, balancing, resilience, and wellbeing.

Compared with MATA, they showed weaker differentiation between related but analytically distinct processes, such as loss of control, adaptation, relational strain, and health-related anxiety. The counterfactual analyses strengthen this interpretation. Across gender, age, and SES, changing only the demographic label reduced lexical overlap by around 30%, indicating that the LLM systematically changed the vocabulary it used when the same underlying topic content was attributed to a different group. Although broader frequency distributions remained similar, the shift in lexical selection suggests that demographic labels are not treated as neutral descriptors.

The sentiment analyses further showed that these lexical shifts were socially patterned rather than random. Higher SES and older age labels produced more positive sentiment, whereas lower SES and male labels produced more negative sentiment. NRC analysis suggested that these shifts were carried by specific emotional vocabularies, including more fear and disgust language for lower SES and lower joy and trust for male-labelled outputs.

This study contributes to emerging work on the use of AI in qualitative analysis, particularly in how socio-demographic variation is represented in health-related free-text data [1,2].

Existing research has largely focused on identifying demographic differences in health outcomes at the population level [4,5], as well as on whether topic modelling and LLMs approaches can capture the complexity and nuance of lived experiences when applied to large volume of free-text data [20,25]. While recent studies have begun to examine how transparent LLM-generated interpretations are, and to address bias in LLM-assisted analysis, they typically focus on output quality or agreement with human coding, rather than how different analytic approaches shape the representation of socio-demographic groups within model outputs [22,23]. Counterfactual testing of whether LLM outputs shift systematically in response to demographic labelling remains uncommon. There is also limited work comparing LLM-based interpretation with established approaches such as MATA and human-led qualitative analysis within a structured evaluation framework [6,30].

Through a systematic evaluation using the GRACE framework, we identified recurring patterns in the LLM outputs that we characterise as “LLM speak”. We use the term “LLM speak” to refer to recurrent “features” in model-generated analysis of qualitative data that create an appearance of coherence and insight while reducing conceptual, contextual and experiential quality of output (Table 8).

**Table 8:** Patterns of “LLM speak” identified in the analysis.

| Pattern of LLM speak | What it means | Illustrative example |
| --- | --- | --- |
| Abstract filler language | Vague, polished phrasing that signals complexity or insight without specifying the analytic content. | “rich tapestry”; “multifaceted nature” |
| Loose conceptual bundling | Analytically distinct concepts are merged into a single label without clarifying their relationship, producing loose rather than precise categories. | “Health and connection”; “Family dynamics and personal health”; “Health, worry, and the impact of isolation”; “Balancing health, work, and well-being”; “Balancing responsibilities and personal well-being” |
| Headline-level summarisation | Findings are presented as broad, plausible headlines rather than as analytically developed interpretations. | “Challenges, disruption and coping strategies”; “Demands and stressors ... and strategies to cope” |
| Interchangeable use of constructs | Different forms, intensities, contexts, or dimensions of experience are compressed into broad categories, reducing nuance and subgroup-specific differentiation. This can also involve the interchangeable use of related constructs. | Mental health, physical health, and wellbeing are used interchangeably across topic labels and descriptions; repeated use of general categories such as “demands”, “pressures”, or “health impacts” across multiple topics. |
| False binary framing | Experience is framed in either/or terms rather than as co-existing, layered, or contradictory. | “While many participants found solace in their relationships, hobbies, and routines, others struggled with feelings of loneliness ...” |

We encourage researchers to use GRACE to identify these and related patterns, in order to build a cumulative understanding of LLM speak and to support the development of alternative methods for analysing human experience that do not rely on LLM “interpretation” of qualitative data.

### Limitations

First, the age stratification, particularly the 18–49 category, covered a broad developmental and life-stage range. This binary grouping was a deliberate simplification, used as a conservative analytic design, and likely reduced finer-grained differentiation within the age-stratified analysis. However, meaningful age-related variation was still identified through MATA despite this conservative approach. Second, LDA is more transparent and less prone to overfitting than newer embedding-based models, but the resulting topics were at times unclear, overlapping, or repetitive, and required substantial researcher interpretation. Some thematic boundaries were therefore difficult to delineate. Third, complete blinding was not always feasible. Certain content within the responses, such as references to grandchildren, could indirectly signal subgroup identity and may have influenced interpretation. Last, although all researchers brought lived experience relevant to the broader topic area, patient and public involvement was not incorporated into the analytic process. Involving public contributors in the interpretation and evaluation of findings will be an important next step, particularly given the study’s focus on representation and socio-demographic difference.

### Implications

#### Research

It is important to analyse SES differences in large volumes of free-text data linked to socio-demographic variables. This offers a useful bridge between qualitative and quantitative approaches. In this context, MATA was particularly valuable. Even with a conservative and simplified design, this hybrid human-AI approach identified meaningful socio-demographic variation with greater conceptual precision and better retention of nuance, whereas LLM-based interpretation produced more overlapping and generalised outputs and was sensitive to demographic framing.

Future research should therefore prioritise human-led, machine-assisted designs and test finer-grained and intersectional subgroup analyses. This is important not only for identifying differences between groups, but also for making less dominant experiences more visible and improving representation of marginalised and otherwise underserved groups in free-text research. Future work should also use structured evaluation frameworks such as GRACE to assess whether model outputs remain interpretable and nuanced, and to build a cumulative evidence base on where LLM use remains analytically defensible and where human expertise is non-negotiable. Relatedly, identifying recurrent patterns of “LLM speak” may help researchers detect when model-generated interpretation becomes overly general, imprecise, or misleading. In addition, patient and public involvement should be incorporated into the evaluation of such analyses, particularly when working with data from underserved or marginalised groups.

### Implications for practice and policy

For public health, service evaluation, and patient-experience analysis, the main risk is not simply that LLMs may be inaccurate, but that they may systematically reframe the same underlying material depending on the demographic label attached to it. In routine analytical settings, this could distort how needs, difficulties, and resources are characterised across social groups, and in doing so obscure or reproduce existing social inequalities.

LLMs may have a role in rapid summarisation or analytic triage, but they should not be used as stand-alone interpretative tools for socially stratified qualitative data. Qualitative researchers should remain directly involved in the analysis of large-scale free-text data, particularly where findings may inform service design, policy, or resource allocation. Human oversight, explicit bias testing, and transparent audit processes are needed before such systems are used in decision-relevant contexts. Without these safeguards, routine use may not only reduce analytic precision but also reinforce stereotyped accounts of social groups and further marginalise less-heard voices.

## Conclusions

This study addressed an important gap in the emerging literature on AI-assisted qualitative analysis by moving beyond questions of whether LLMs can produce plausible thematic summaries, and instead examining how their outputs perform when socio-demographic sensitivity, interpretative precision, and bias are central to the research question.

Using a combined qualitative and quantitative evaluation, we showed that MATA preserved more meaningful socio-demographic variation, clearer conceptual structure, and greater interpretative depth than LLM-generated “interpretation”, while counterfactual relabelling revealed that LLM outputs were also sensitive to demographic cues in socially patterned ways.

Together, these findings suggest that LLMs are insufficient as standalone interpretative tools for lived-experience free-text data where nuanced understanding and fair representation matter. More broadly, the study offers a systematic approach for evaluating AI-assisted text analysis, combining structured qualitative appraisal with potential bias testing, and underlines why such evaluation is necessary if computational methods are to be used responsibly in human-centred research. This is particularly important when analysing the experiences of marginalised or less-heard groups, whose perspectives may otherwise be flattened, reframed, or lost in automated summaries.

## Data Availability

The data that support the findings of this study are available on reasonable request from the study sponsor: Faculty of Medicine and Health Research and Innovation Services.

## Acknowledgments

We would like to thank Sarah Taylor for her advice and editorial assistance during manuscript preparation.

## Supporting information captions

Appendix 1 - Prompt engineering content used

Appendix 2 - Identification of modifications to the excerpts of the LLM analysis

